# Neutralization Escape by the SARS-CoV-2 Omicron Variants BA.2.12.1 and BA.4/BA.5

**DOI:** 10.1101/2022.05.16.22275151

**Authors:** Nicole P. Hachmann, Jessica Miller, Ai-ris Y. Collier, John D. Ventura, Jingyou Yu, Marjorie Rowe, Esther Apraku Bondzie, Olivia Powers, Nehalee Surve, Kevin Hall, Dan H. Barouch

## Abstract

Multiple lineages of the SARS-CoV-2 Omicron variant (B.1.1.529) have emerged, and BA.1 and BA.2 have demonstrated substantial escape from neutralizing antibodies (NAbs). BA.2.12.1 has now become dominant in the United States, and BA.4 and BA.5 have become dominant in South Africa. Our data show that BA.2.12.1 and BA.4/BA.5 substantially escape NAbs induced by both vaccination and infection. Moreover, BA.4/BA.5 NAb titers, and to lesser extent BA.2.12.1 NAb titers, were lower than BA.1 and BA.2 NAb titers, suggesting that the SARS-CoV-2 Omicron variant has continued to evolve with increasing neutralization escape. These findings have important public health implications and provide immunologic context for the current surges with BA.2.12.1 and BA.4/BA.5 in populations with high rates of vaccination and BA.1/BA.2 infection.

---

Multiple lineages of the SARS-CoV-2 Omicron variant (B.1.1.529) have emerged^1^, and BA.1 and BA.2 have demonstrated substantial escape from neutralizing antibodies (NAbs)^2-5^. BA.2.12.1 has now become dominant in the United States, and BA.4 and BA.5 have become dominant in South Africa (**Fig. 1A**). BA.4 and BA.5 have identical Spike sequences. The ability of BA.2.12.1 and BA.4/BA.5 to evade NAbs induced by vaccination or infection has not yet been reported.

**Figure 1.**
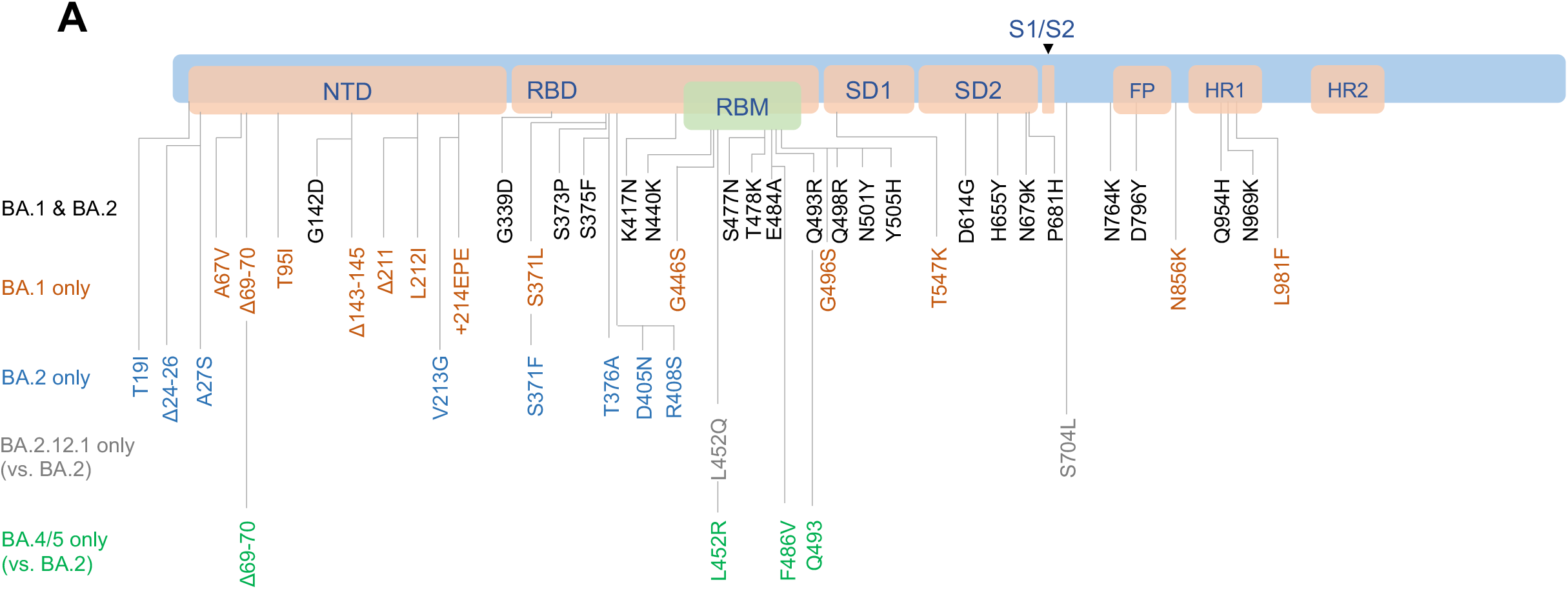
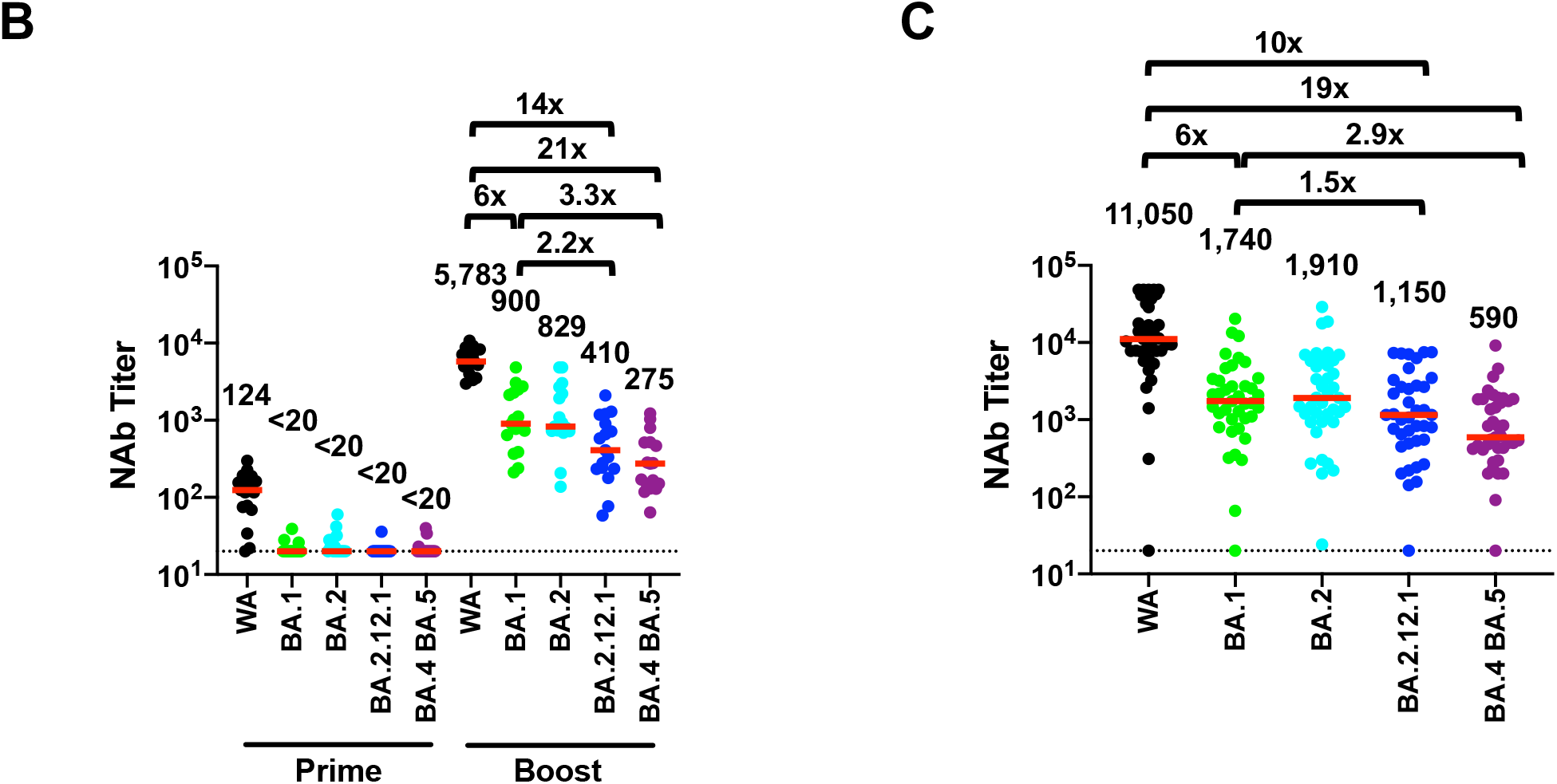
Neutralizing antibody responses to Omicron subvariants. **A**. Cartoon showing BA.1, BA.2, BA.2.12.1, BA.4, and BA.5 mutations in the SARS-CoV-2 Spike. BA.4 and BA.5 have identical Spike sequences. NTD, N-terminal domain; RBD, receptor binding domain; RBM, receptor binding motif; SD1, subdomain 1; SD2, subdomain 2; FP, fusion peptide; HR1, heptad repeat 1; HR2, heptad repeat 2. **B**. Neutralizing antibody (NAb) titers by a luciferase-based pseudovirus neutralization assay in individuals six months following initial BNT162b2 vaccination (Prime) and two weeks following BNT162b2 boost (Boost). **C**. NAb titers in individuals following infection with BA.1 or BA.2. All were vaccinated except for the one individual with negative NAb titers. NAb responses were measured against the SARS-CoV-2 WA1/2020, Omicron BA.1, BA.2, BA.2.12.1, and BA.4/BA.5 variants. Medians (red bars) are depicted and shown numerically with fold differences.

We evaluated WA1/2020, Omicron BA.1, BA.2, BA.2.12.1, and BA.4/BA.5 NAb titers in 27 individuals who were vaccinated and boosted with the mRNA vaccine BNT162b2 and in 27 individuals who were infected with SARS-CoV-2 Omicron BA.1 or BA.2 (**Table S1**). In the vaccine cohort, individuals were excluded if they had a history of SARS-CoV-2 infection or a positive nucleocapsid (N) serology, or if they received other COVID-19 vaccines or immunosuppressive medications. Six months after the initial two BNT162b2 immunizations, median pseudovirus NAb responses were 124 to WA1/2020 but <20 to all Omicron subvariants (**Fig. 1B**). Two weeks following the boost, median NAb titers increased substantially to 5783, 900, 829, 410, and 275 to WA1/2020, Omicron BA.1, BA.2, BA.2.12.1, and BA.4/BA.5, respectively. These data show a 6.4-, 7.0-, 14.1-, and 21.0-fold reduction of median NAb titers to BA.1, BA.2, BA.2.12.1, and BA.4/BA.5 compared with WA1/2020, respectively. Median BA.2.12.1 NAb titers were 2.2-fold lower and median BA.4/BA.5 NAb titers were 3.3-fold lower than median BA.1 NAb titers.

In the Omicron infection cohort, all but one individual were vaccinated, and samples were obtained a median of 29 days following diagnosis (**Table S2**). Median NAb titers were 11,050, 1740, 1910, 1150, and 590 to WA1/2020, Omicron BA.1, BA.2, BA.2.12.1, and BA.4/BA.5, respectively (**Fig. 1C**). These data show a 6.4-, 5.8-, 9.6-, and 18.7-fold reduction of median NAb titers to BA.1, BA.2, BA.2.12.1, and BA.4/BA.5 compared with WA1/2020, respectively. Median BA.2.12.1 NAb titers were 1.5-fold lower and median BA.4/BA.5 NAb titers were 2.9-fold lower than median BA.1 NAb titers.

These data show that BA.2.12.1 and BA.4/BA.5 substantially escape NAbs induced by both vaccination and infection. Moreover, BA.4/BA.5 NAb titers, and to lesser extent BA.2.12.1 NAb titers, were lower than BA.1 and BA.2 NAb titers, suggesting that the SARS-CoV-2 Omicron variant has continued to evolve with increasing neutralization escape. These findings have important public health implications and provide immunologic context for the current surges with BA.2.12.1 and BA.4/BA.5 in populations with high rates of vaccination and BA.1/BA.2 infection.

## Data sharing

N.P.H. and D.H.B. had full access to all the data in the study and take responsibility for the integrity of the data and the accuracy of the data analysis. All data are available in the manuscript or the supplementary material. Correspondence and requests for materials should be addressed to D.H.B.

## Data Availability

All data produced in the present work are contained in the manuscript.

## Funding

The authors acknowledge NIH grant CA260476, the Massachusetts Consortium for Pathogen Readiness, the Ragon Institute, and the Musk Foundation (D.H.B.), as well as the Reproductive Scientist Development Program from the Eunice Kennedy Shriver National Institute of Child Health & Human Development and Burroughs Wellcome Fund HD000849 and NIH grant AI69309 (A.Y.C.).

### Role of Sponsor

The sponsor did not have any role in design or conduct of the study; collection, management, analysis, or interpretation of the data; preparation, review, or approval of the manuscript; or decision to submit the manuscript for publication.

### Conflicts of Interest

The authors report no conflicts of interest.

### Author Contributions

D.H.B.: conceptualization, formal analysis, resources, investigation, data curation, writing-original draft, writing-review & editing, visualization, supervision

N.P.H., J.M., A.Y.C., J.D.V., J.Y., M.R., E.A.B., O.P., N.H., K.H.: investigation, methodology, data curation, writing-review & editing

## Acknowledgements

The authors thank the study participants and the Center for Virology and Vaccine Research Clinical Trials Unit for enrollment and processing samples for the BIDMC COVID-19 Biorepository.

## Supplementary Methods

### Study population

A specimen biorepository at Beth Israel Deaconess Medical Center (BIDMC) obtained samples from individuals who received a SARS-CoV-2 vaccine and/or had a history of SARS-CoV-2 infection. The BIDMC institutional review board approved this study (2020P000361). All participants provided informed consent. For the group of individuals receiving three doses of BNT162b2, participants were excluded if they had a history of SARS-CoV-2 infection or a positive nucleocapsid (N) serology by electrochemiluminescence assays (ECLA), or if they received other COVID-19 vaccines or immunosuppressive medications.

### Pseudovirus neutralizing antibody assay

The SARS-CoV-2 pseudoviruses expressing a luciferase reporter gene were used to measure pseudovirus neutralizing antibodies. In brief, the packaging construct psPAX2 (AIDS Resource and Reagent Program), luciferase reporter plasmid pLenti-CMV Puro-Luc (Addgene) and spike protein expressing pcDNA3.1-SARS-CoV-2 SΔCT were co-transfected into HEK293T cells (ATCC CRL_3216) with lipofectamine 2000 (ThermoFisher Scientific). Pseudoviruses of SARS-CoV-2 variants were generated by using WA1/2020 strain (Wuhan/WIV04/2019, GISAID accession ID: EPI_ISL_402124), Omicron B.1.1.529 BA.1 (GISAID ID: EPI_ISL_7358094.2), or BA.2 (GISAID ID: EPI_ISL_6795834.2), BA.2.12.1 (GISAID ID: EPI_ISL_12003853.1), and BA.4/BA.5 (GSAID ID: EPI_ISL_12268495.2). The supernatants containing the pseudotype viruses were collected 48h after transfection; pseudotype viruses were purified by filtration with 0.45-μm filter. To determine the neutralization activity of human serum, HEK293T-hACE2 cells were seeded in 96-well tissue culture plates at a density of 2 × 10^4^ cells per well overnight. Three-fold serial dilutions of heat-inactivated serum samples were prepared and mixed with 50 μl of pseudovirus. The mixture was incubated at 37 °C for 1 h before adding to HEK293T-hACE2 cells. After 48 h, cells were lysed in Steady-Glo Luciferase Assay (Promega) according to the manufacturer’s instructions. SARS-CoV-2 neutralization titers were defined as the sample dilution at which a 50% reduction (NT50) in relative light units was observed relative to the average of the virus control wells.

**Table S1.** Study Population.

|  | **BNT162b2**<br>N=27 | **Omicron infection**<br>N=27 |
| --- | --- | --- |
| **Age (years), median (range)** | 35 (23-76) | 34 (27-41) |
| **Sex at birth, female** | 24 (89) | 21 (78) |
| **Race** |  |  |
| White | 23 (85) | 21 (78) |
| Asian | 2 (7) | 1 (4) |
| Black | 1 (4) | 2 (7) |
| More than one race | 1 (4) | 0 |
| Unknown/Other* | 0 (0) | 3 (11) |
| **Ethnicity** |  |  |
| Hispanic or Latino | 2 (7) | 1 (4) |
| Non-Hispanic | 24 (89) | 24 (89) |
| Other | 1 (4) | 2 (7) |
| **Medical condition** |  |  |
| Obese (BMI $\geq 30$ kg/m<sup>2</sup>) | 2 (7) | 2 (7) |
| Hypertension | 3 (11) | 4 (15) |
| Diabetes | 2 (7) | 3 (11) |
| Pregnant | 2 (7) | 7 (26) |
| Lactating | 4 (15) | 3 (11) |
| Asthma | 0 | 3 (11) |
| **COVID-19 vaccination history** |  |  |
| BNT162b2 (3 doses) | 27 (100) | 8 (30) |
| BNT162b2 (2 doses)/mRNA-1273 (1 dose) | 0 | 2 (7) |
| mRNA-1273 (3 doses) | 0 | 7 (26) |
| mRNA-1273 (2 doses)/BNT-1273 (1 dose) | 0 | 2 (7) |
| mRNA-1273 (2 doses)/Ad26.COV2.S (1 dose) | 0 | 1 (4) |
| BNT162b2 (2 doses) | 0 | 4 (15) |
| Ad26.COV2.S (1 dose)/mRNA-1273 (1 dose) | 0 | 1 (4) |
| Ad5/Ad26 (2 doses)/mRNA-1273 (1 dose) | 0 | 1 (4) |
| Unvaccinated | 0 | 1 (4) |
| **COVID-19 disease severity (NIH scale)** |  |  |
| Asymptomatic | N/A | 1 (4) |
| Mild | N/A | 24 (89) |
| Moderate | N/A | 0 |
| Severe | N/A | 2 (7) |
| Critical | N/A | 0 |
| **Days from last vaccine dose to positive PCR test, median (IQR)** | N/A | 83 (37-153) |
| **Days from positive PCR test to sampling, median (IQR)** | N/A | 29 (14-74) |

Data displayed as median (range or interquartile range, IQR) and n (%). BMI, body mass index; NIH, National Institutes of Health; PCR, polymerase chain reaction; pregnant designation reflects time of sampling. \*Self-identified race as Jamaican, Brazilian, and Colombian.

**Table S2.** SARS-CoV-2 Infected Participants.

| ID | Dominant variant* | Medical history | COVID-19 vaccine history (primary/boost) | Days from boost or last dose to PCR | Days from PCR to sampling | COVID-19 NIH disease severity |
| --- | --- | --- | --- | --- | --- | --- |
| 1 | BA.1 |  | BNT162b2 | 69 | 106 | mild |
| 2 | BA.1 |  | BNT162b2/<br>mRNA-1273 | 26 | 81 | mild |
| 3 | BA.1 | Asthma | BNT162b2/<br>mRNA-1273 | 40 | 74 | mild |
| 4 | BA.1 | Pregnant | mRNA-1273/<br>mRNA-1273 | 59 | 52, 102 | mild |
| 5 | BA.1 |  | mRNA-1273/<br>mRNA-1273 | 67 | 13, 113 | mild |
| 6 | BA.1 | Pregnant | mRNA-1273/<br>mRNA-1273 | 21 | 12 | mild |
| 7 | BA.1 | HTN | BNT162b2/<br>BNT162b2 | 88 | 25, 109 | mild |
| 8 | BA.1 | Pregnant, Obesity | BNT162b2 | 287 | 17 | mild |
| 9 | BA.1 | Obesity | Ad26.COV2.S/<br>mRNA-1273 | 11 | 15, 102 | mild |
| 10 | BA.1 |  | BNT162b2/<br>BNT162b2 | 99 | 22 | mild |
| 11 | BA.1 |  | mRNA-1273/<br>Ad26.COV2.S | 67 | 51, 94 | mild |
| 12 | BA.1 | DM, HTN | mRNA-1273/<br>mRNA-1273 | 6 | 8 | asymptomatic |
| 13 | BA.1 | DM | BNT162b2 | 210 | 9 | severe |
| 14 | BA.1 | Pregnant | Unvaccinated | N/A | 4 | severe |
| 15 | BA.1 | Pregnant | mRNA-1273/<br>mRNA-1273 | 77 | 35 | mild |
| 16 | BA.1 | Pregnant | BNT162b2 | 228 | 2 | mild |
| 17 | BA.1 | Asthma | BNT162b2/<br>BNT162b2 | 101 | 13, 71 | mild |
| 18 | BA.1 | Asthma | mRNA-1273/<br>BNT162b2 | 15 | 14 | mild |
| 19 | BA.1 |  | mRNA-1273/<br>mRNA-1273 | 11 | 33 | mild |
| 20 | BA.1 | Lactating | BNT162b2/<br>BNT162b2 | 116 | 10, 54 | mild |
| 21 | BA.1 |  | mRNA-1273/<br>BNT162b2 | 42 | 16, 29, 91 | mild |
| 22 | BA.1 | Lactating, HTN | BNT162b2/<br>BNT162b2 | 102 | 18, 74 | mild |
| 23 | BA.1 | Pregnant | Sputnik V<br>(Ad5/Ad26)/<br>mRNA-1273 | 239 | 32 | mild |
| 24 | BA.1 | Lactating | BNT162b2/<br>BNT162b2 | 113 | 41 | mild |
| 25 | BA.1 | DM | BNT162b2/<br>BNT162b2 | 164 | 12 | mild |
| 26 | BA.2 | HTN | BNT162b2/<br>BNT162b2 | 169 | 16 | mild |
| 27 | BA.2 |  | mRNA-1273/<br>mRNA-1273 | 149 | 16 | mild |

\* >50% of sequences in New England (Region 1) according to CDC; PCR, polymerase chain reaction for SARS-CoV-2; NIH, National Institutes of Health; HTN, hypertension; DM, diabetes mellitus.

## Notes

### Competing Interest Statement

The authors have declared no competing interest.

### Author Declarations

IRB of Beth Israel Deaconess Medical Center gave ethical approval for this work.

